# Real-World Use, Dosing, and Reported Side Effects of BPC-157 and TB-500 for Musculoskeletal Injury: An Analysis of Online Communities

**DOI:** 10.64898/2026.11.04.26367022

**Authors:** Joshua Cassinat, Claire Callan, Catherine Liggett, Vonda Wright

## Abstract

**Objective:** To describe how people use the unapproved peptides BPC-157 and TB-500 for tendon, ligament and muscle injury and what side effects they report.

**Methods:** Using a large language model validated against physician review, we analyzed 92,254 posts and comments from 28,652 users in Reddit’s peptide communities from 2021 to 2026. We compared side effects with those reported by users of four other peptides in the same communities.

**Results:** In all, 5,588 users described using BPC-157 and/or TB-500 for a specific musculoskeletal injury, most often a chronic tendon problem of the shoulder, knee or elbow. Only 3.8% of users described physician involvement, and 90.9% of those naming a source bought from research-chemical vendors. The median BPC-157 dose (366 µg per injection) was about 2.7 times the human-equivalent dose of the 10 µg/kg rat regimen, and among new users it rose from 300 µg in 2024 to 500 µg in 2025. Of those stating an outcome, 79.3% reported improvement, and 36.6% of all users were also in physical therapy. Overall, 27.2% described at least one side effect, most often fatigue, anxiety, anhedonia and insomnia. Anhedonia was more common than among users of the four other peptides (10.2% versus 1.3% of those with side effects).

**Conclusion:** Use of these unapproved peptides for musculoskeletal injury is largely unsupervised, relying on grey-market products at self-selected, rising doses without established safety or efficacy.

**What is already known on this topic:**

- Evidence that BPC-157 and TB-500 help tendon, ligament or muscle injury comes almost entirely from animal studies.
- Both are prohibited in sport by the World Anti-Doping Agency, and in July 2026 a US Food and Drug Administration advisory committee voted to recommend them for pharmacy compounding.

**What this study adds:**

- We analyzed over 90,000 Reddit posts from 5,588 users taking these peptides for a specific injury, most often a chronic tendon problem of the shoulder, knee or elbow.
- Only 3.8% described physician involvement, and 90.9% of those naming a source bought from research-chemical vendors.
- The median BPC-157 dose was about 2.7 times the human-equivalent dose of the rat regimen, and 27.2% of users reported a side effect.
- Anhedonia was about eight times as common as among users of four other peptides in the same communities.

**How this study might affect research, practice or policy:**

- Clinicians treating chronic tendon problems should ask about peptide use and counsel patients on the lack of human efficacy data, unregulated supply, anti-doping status and reported side effects.
- Trials with prespecified neuropsychiatric outcomes are needed before any benefit or dose can be claimed.
- These findings may inform regulatory decisions on pharmacy compounding.

## Introduction

Two unapproved peptides, BPC-157 and TB-500, have rapidly gained popularity as “healing peptides” for tendon, ligament and muscle injury.^1–4^ BPC-157, a synthetic 15–amino acid fragment of a protein isolated from human gastric juice, promotes the survival and migration of tendon fibroblasts and increases their growth hormone receptor expression in cultured rat cells.^5,6^ In rats, it accelerated healing of transected or detached Achilles tendons, transected ligaments and injured myotendinous junctions, in studies almost entirely from a single research group.^7–10^ Its half-life in rats and dogs is under 30 minutes.^11^ Human data are limited to a retrospective series in which 7 of 12 patients reported relief of knee pain after intra-articular injection^12^ and a small intravenous safety pilot,^13^ and a 2025 systematic review found 35 preclinical studies, one clinical study and no controlled trials or clinical safety data.^14^

TB-500 is a synthetic version of the acetylated 17–23 fragment of thymosin β4,^15^ an actin-sequestering protein that promotes wound repair in animal models.^16^ Full-length thymosin β4 has been tested in clinical trials of eye and skin wound healing,^17^ but we found no published clinical study of the TB-500 fragment.

Despite this popularity, there are no systematic data on how these peptides are used, and no study has established an effective or safe human dose. Products are sold online as research chemicals,^1,4^ so their use is unlikely to be captured by health records or spontaneous adverse event reporting. Both peptides are prohibited at all times by the World Anti-Doping Agency,^18^ yet in July 2026 a US Food and Drug Administration (FDA) advisory committee voted to recommend both for pharmacy compounding.^19^ Clinicians, athletes and regulators therefore have little information on how these peptides are used or what harm they cause.

Online health communities are one source of such data: users describe their own treatment experiences in detail and without prompting, and analyses of such posts can detect side effects under-recognized in trials and post-marketing surveillance.^20^ Large language models now allow such posts to be analyzed at scale. Using this approach, Sehgal and colleagues described self-reported side effects among more than 67,000 Reddit users of semaglutide and tirzepatide, identifying menstrual and temperature-related symptoms not recognized in drug labeling,^21^ and then studied retatrutide.^22^ Similar analyses have described side effects of grey-market selective androgen receptor modulators.^23^

We therefore analyzed 92,254 posts and comments from Reddit’s peptide communities with three objectives: to describe (1) the injuries treated, regimens, sources, physician involvement and reported outcomes of people using BPC-157 and TB-500 for musculoskeletal injury, and (2) the side effects they attribute to these peptides; and (3) to test whether any side effect is specific to BPC-157 and TB-500 rather than a feature of how these communities report side effects, by comparing their users with users of four other peptides discussed in the same communities.

## Methods

### Data source

We collected all posts and comments from 1 January 2021 to 31 August 2026 in the six subreddits dedicated to peptide use (r/Peptides, r/bpc_157, r/peptidesource, r/PeptideGuide, r/TB500 and r/Semax) from Arctic Shift, a public archive of Reddit data, and kept those mentioning BPC-157, TB-500 or four other peptides considered by the FDA advisory committee (Supplementary Appendix). Following prior work,^21^ we excluded deleted and removed content, bot accounts, texts under 10 words and texts with fewer than 50% alphabetic characters, leaving 92,254 posts and comments from 28,652 users (6.6 million words). We followed the Strengthening the Reporting of Observational Studies in Epidemiology (STROBE) guidelines.

### Language-model extraction

We used a large language model (Qwen3-235B-A22B-Instruct-2507) to identify posts describing the author’s own use of a study peptide and, for BPC-157 and TB-500 users describing a musculoskeletal problem, to extract the injury’s location, tissue and chronicity, the dose, route and source, concurrent treatments, physician involvement and the reported outcome. Each side effect the author attributed to the peptide was mapped to a Medical Dictionary for Regulatory Activities (MedDRA) Preferred Term (PT) and system organ class (SOC) and classed as serious or non-serious using the regulatory definition. Further details are in the Supplementary Appendix.

### Validation

Physicians blinded to the model’s output annotated 400 randomly selected posts. Against these annotations, the model identified musculoskeletal use with a sensitivity of 94% and a precision of 96% (κ=0.90), physician involvement with κ=0.89, the reported outcome with κ=0.67, self-use with a sensitivity of 75% and a precision of 91%, and posts describing a side effect with a sensitivity of 85% and a precision of 79%. A physician reviewed every event the model flagged as serious.

### Statistical analysis

Side-effect percentages were calculated among users reporting at least one side effect, as in prior work,^21^ and percentages for injuries, injection site and source among users who stated each item, with 95% Wilson confidence intervals. Sensitivity analyses excluded injection-site terms, excluded users reporting co-use of incretin-based drugs, androgens or selective androgen receptor modulators, growth hormone or its secretagogues, or stimulants, and examined BPC-157 with and without TB-500. As a negative control, we compared users who reported only BPC-157 and/or TB-500 with users who reported only one of the other four peptides, using risk ratios among users reporting side effects and odds ratios adjusted for posting volume. Physician-involved and other users, and users taking 500 µg or more of BPC-157 per injection and other users, were compared by logistic regression adjusted for posting volume and, for dose, year of first post. A user was counted as having had surgery if either the injury or the treatment extraction said so, and growth in new users was compared with the number of authors posting about any study peptide each year. Analyses used Python 3.13.

### Equity, diversity and inclusion statement

The author team includes senior and early career researchers from diverse medical specialties, including orthopaedics and rehabilitation medicine, and is majority female-led. We did not extract race, ethnicity or income. Fewer than one in ten users stated their age or sex, and those who did were mostly male, so our findings may not apply equally to women, older adults or people who do not post in English. We report only aggregate data.

### Patient and public involvement

Patients and the public were not involved in the design, conduct or reporting of this study.

## Results

The model classified all 92,254 posts and comments; 34,071 described personal use by 13,456 users. Of these, 6,281 described using BPC-157 and/or TB-500 for a musculoskeletal problem, and the 5,588 who named a specific body region or tissue formed the cohort (Figure 1). Where stated, users were mostly male (69.7% of 373), with a median age of 42 years (interquartile range [IQR] 33–50; n=455).

**Figure 1.**
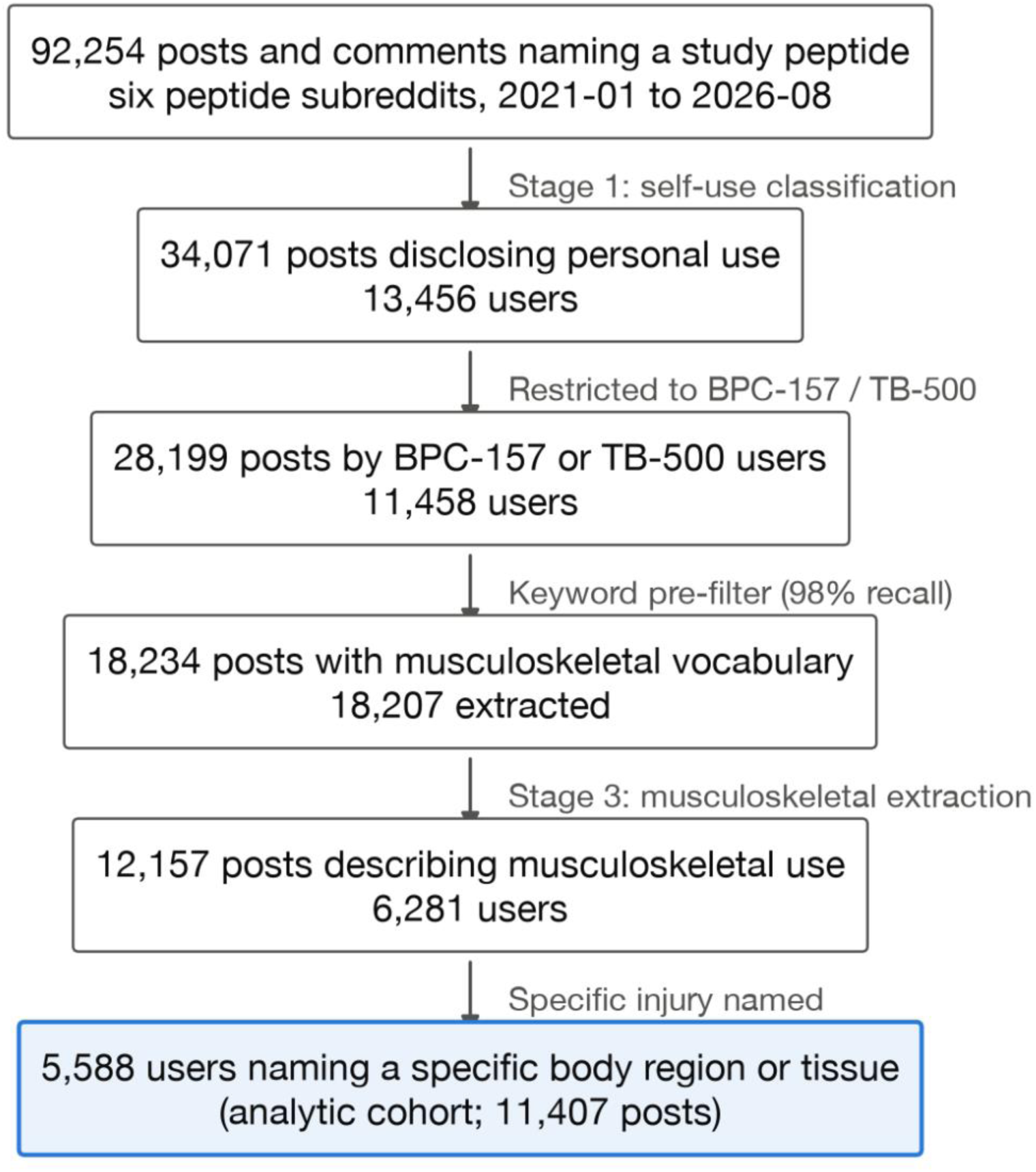
Study flow. Counts are texts (posts and comments) unless stated as users.

### Injuries and regimens

Injuries were mostly chronic (75.2%) and tendinous (50.4%), most often of the shoulder (38.9%), knee (24.8%), elbow (18.4%) and lumbar or thoracic spine (17.6%) (Table 1; Figure 2). Over a third of users (36.6%) were also receiving physical therapy, and 17.4% had undergone surgery for the same problem. The median BPC-157 dose was 366 µg per injection (IQR 250– 500), or about 4.3 µg/kg for an 85 kg adult: less than half the 10 µg/kg regimen of rat tendon and ligament studies,^7–10^ which only 9.0% reached (Figure 3), but about 2.7 times its human-equivalent dose of 1.6 µg/kg by body-surface-area conversion, which 98.0% met or exceeded.^24^ Most users injected subcutaneously (76.3%) and, where a site was stated, away from the injury (86.4%). Of users naming a source, 90.9% bought from research-chemical vendors (90.8% excluding r/peptidesource, a vendor-review community). Physician prescribing or administration was described by only 3.8% of all users, including 5.9% of those who had undergone surgery and 6.4% of those receiving physical therapy.

**Figure 2.**
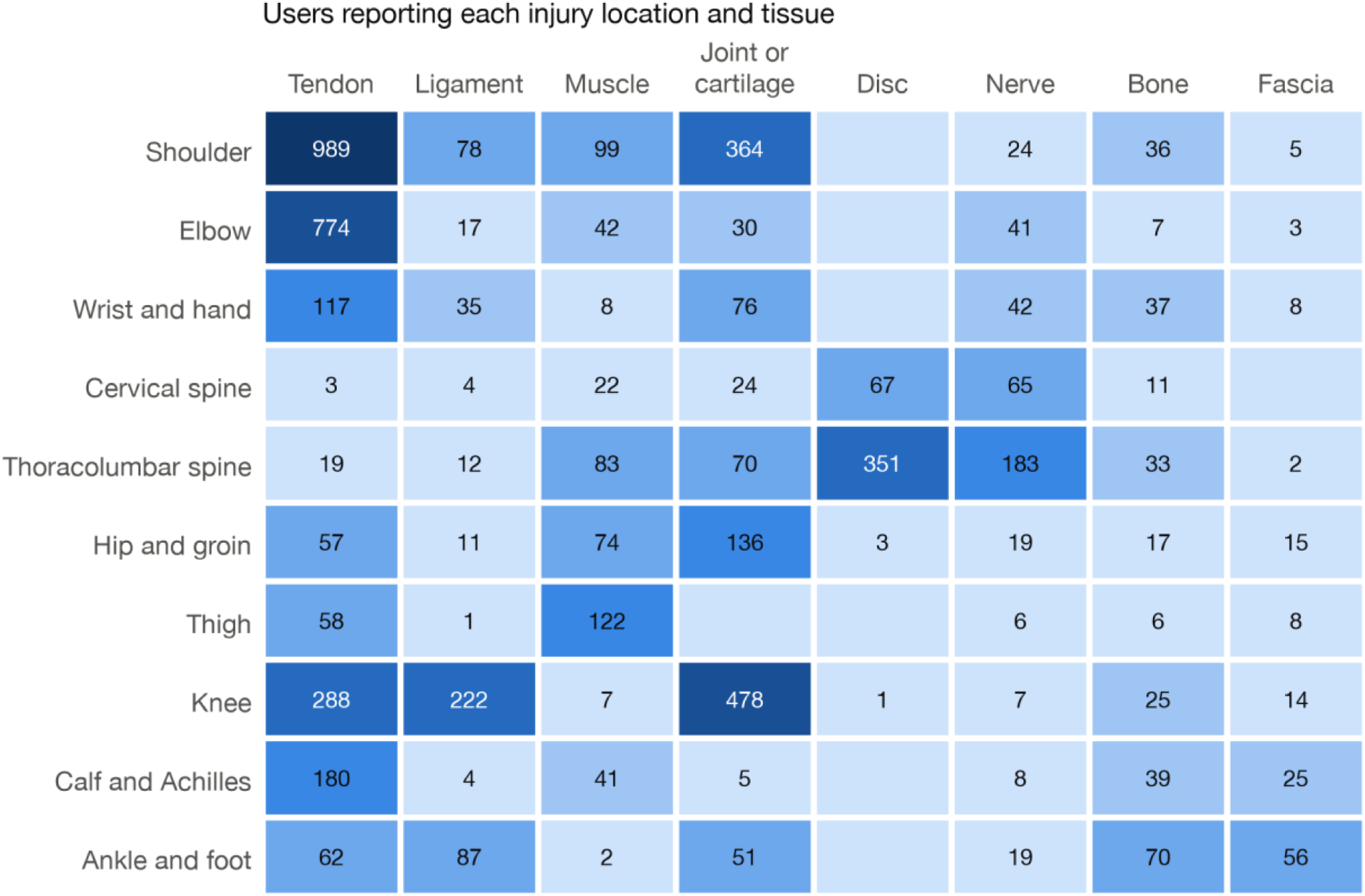
Users reporting each combination of injury region and tissue. A user may contribute more than one cell.

**Figure 3.**
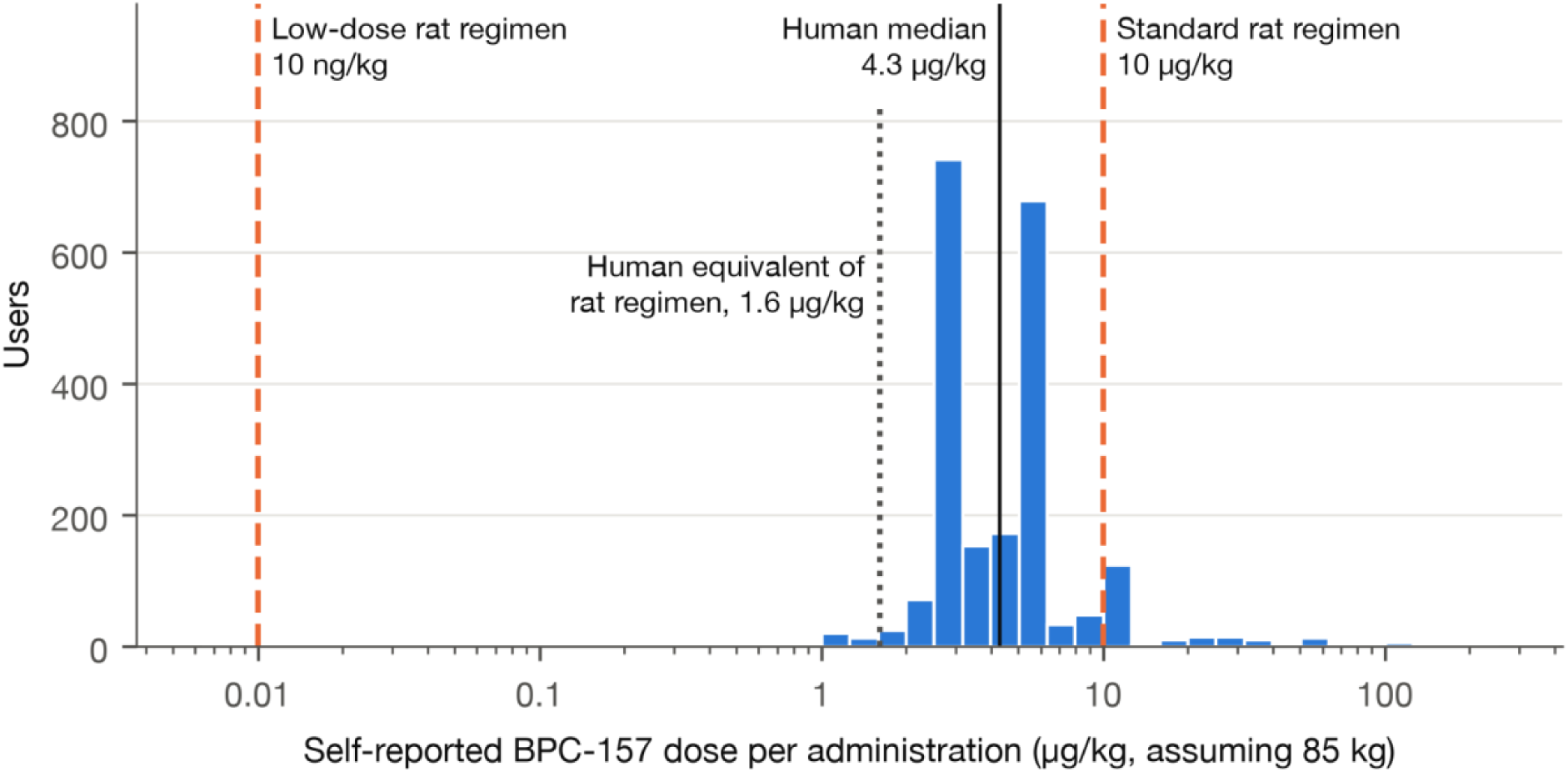
Self-reported BPC-157 dose per injection, expressed per kilogram assuming 85 kg body weight (n=2,158 users), against the two rat regimens used in the musculoskeletal studies (10 µg/kg and 10 ng/kg).^7–10^ The dotted line marks the human-equivalent dose of the 10 µg/kg rat regimen by body-surface-area conversion (1.6 µg/kg).^24^ Logarithmic scale.

**Table 1.**
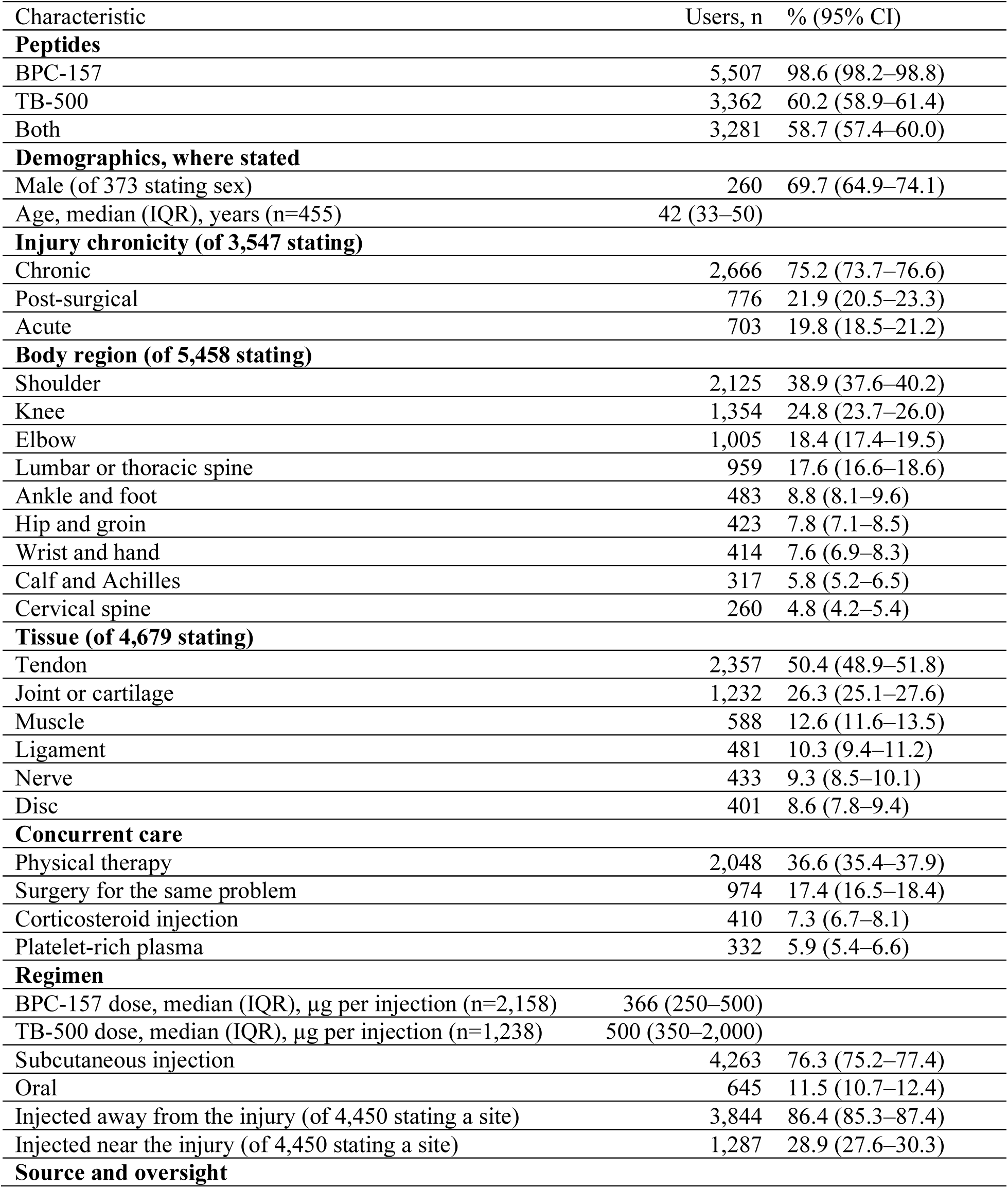
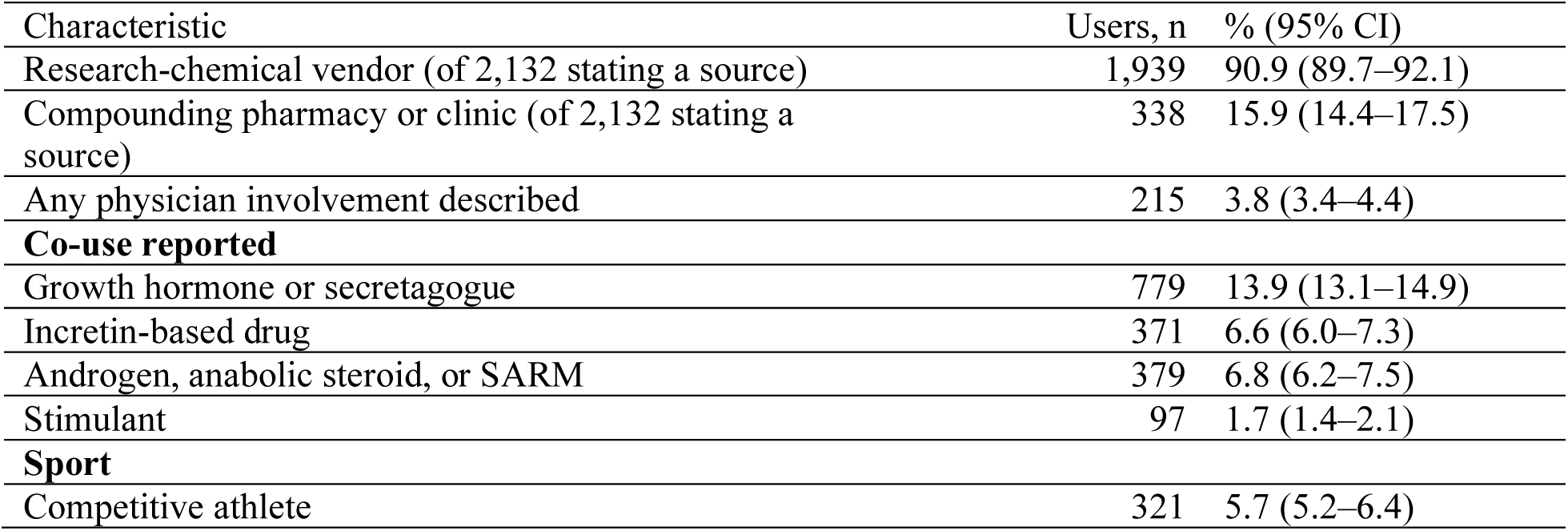
Characteristics, injuries, and regimens of 5,588 Reddit users describing musculoskeletal use of BPC-157 and/or TB-500. Categories are not mutually exclusive. Injury, injection-site, and source percentages use users who stated the item as the denominator.

| Characteristic | Users, n | % (95% CI) |
|---|---|---|
| **Peptides** |  |  |
| BPC-157 | 5,507 | 98.6 (98.2–98.8) |
| TB-500 | 3,362 | 60.2 (58.9–61.4) |
| Both | 3,281 | 58.7 (57.4–60.0) |
| **Demographics, where stated** |  |  |
| Male (of 373 stating sex) | 260 | 69.7 (64.9–74.1) |
| Age, median (IQR), years (n=455) | 42 (33–50) |  |
| **Injury chronicity (of 3,547 stating)** |  |  |
| Chronic | 2,666 | 75.2 (73.7–76.6) |
| Post-surgical | 776 | 21.9 (20.5–23.3) |
| Acute | 703 | 19.8 (18.5–21.2) |
| **Body region (of 5,458 stating)** |  |  |
| Shoulder | 2,125 | 38.9 (37.6–40.2) |
| Knee | 1,354 | 24.8 (23.7–26.0) |
| Elbow | 1,005 | 18.4 (17.4–19.5) |
| Lumbar or thoracic spine | 959 | 17.6 (16.6–18.6) |
| Ankle and foot | 483 | 8.8 (8.1–9.6) |
| Hip and groin | 423 | 7.8 (7.1–8.5) |
| Wrist and hand | 414 | 7.6 (6.9–8.3) |
| Calf and Achilles | 317 | 5.8 (5.2–6.5) |
| Cervical spine | 260 | 4.8 (4.2–5.4) |
| **Tissue (of 4,679 stating)** |  |  |
| Tendon | 2,357 | 50.4 (48.9–51.8) |
| Joint or cartilage | 1,232 | 26.3 (25.1–27.6) |
| Muscle | 588 | 12.6 (11.6–13.5) |
| Ligament | 481 | 10.3 (9.4–11.2) |
| Nerve | 433 | 9.3 (8.5–10.1) |
| Disc | 401 | 8.6 (7.8–9.4) |
| **Concurrent care** |  |  |
| Physical therapy | 2,048 | 36.6 (35.4–37.9) |
| Surgery for the same problem | 974 | 17.4 (16.5–18.4) |
| Corticosteroid injection | 410 | 7.3 (6.7–8.1) |
| Platelet-rich plasma | 332 | 5.9 (5.4–6.6) |
| **Regimen** |  |  |
| BPC-157 dose, median (IQR), µg per injection (n=2,158) | 366 (250–500) |  |
| TB-500 dose, median (IQR), µg per injection (n=1,238) | 500 (350–2,000) |  |
| Subcutaneous injection | 4,263 | 76.3 (75.2–77.4) |
| Oral | 645 | 11.5 (10.7–12.4) |
| Injected away from the injury (of 4,450 stating a site) | 3,844 | 86.4 (85.3–87.4) |
| Injected near the injury (of 4,450 stating a site) | 1,287 | 28.9 (27.6–30.3) |
| **Source and oversight** |  |  |
| Characteristic | Users, n | % (95% CI) |
| Research-chemical vendor (of 2,132 stating a source) | 1,939 | 90.9 (89.7–92.1) |
| Compounding pharmacy or clinic (of 2,132 stating a source) | 338 | 15.9 (14.4–17.5) |
| Any physician involvement described | 215 | 3.8 (3.4–4.4) |
| **Co-use reported** |  |  |
| Growth hormone or secretagogue | 779 | 13.9 (13.1–14.9) |
| Incretin-based drug | 371 | 6.6 (6.0–7.3) |
| Androgen, anabolic steroid, or SARM | 379 | 6.8 (6.2–7.5) |
| Stimulant | 97 | 1.7 (1.4–2.1) |
| **Sport** |  |  |
| Competitive athlete | 321 | 5.7 (5.2–6.4) |

### Reported outcomes

Among 4,832 users stating an outcome, 79.3% reported improvement, 12.7% no change, 4.0% worsening, 2.0% that it was too early to tell and 1.9% mixed results. The median time to a perceived effect was 10.5 days (IQR 7–21).

### Side effects

Overall, 1,519 users (27.2%) reported at least one side effect, with a mean of 2.8 PTs each (Table 2). Among them, the most common were fatigue (13.2%), anxiety (12.8%), anhedonia (11.0%), insomnia (10.1%) and injection site pain (9.8%). A psychiatric symptom was reported by 33.2% and a nervous system symptom by 28.3%, compared with 15.4% for gastrointestinal symptoms. The model flagged 173 potentially serious events from 59 users. On physician review, 29 users (0.5%) described an event meeting the regulatory definition of serious, most often a new or progressing cancer (6 users, 4 with basal cell carcinoma), a persistent or disabling illness (5) or a cardiovascular emergency (5) (Supplementary Table S12); excluding basal cell carcinoma, which not every definition counts as serious, 25 users (0.4%) did.

**Table 2.**
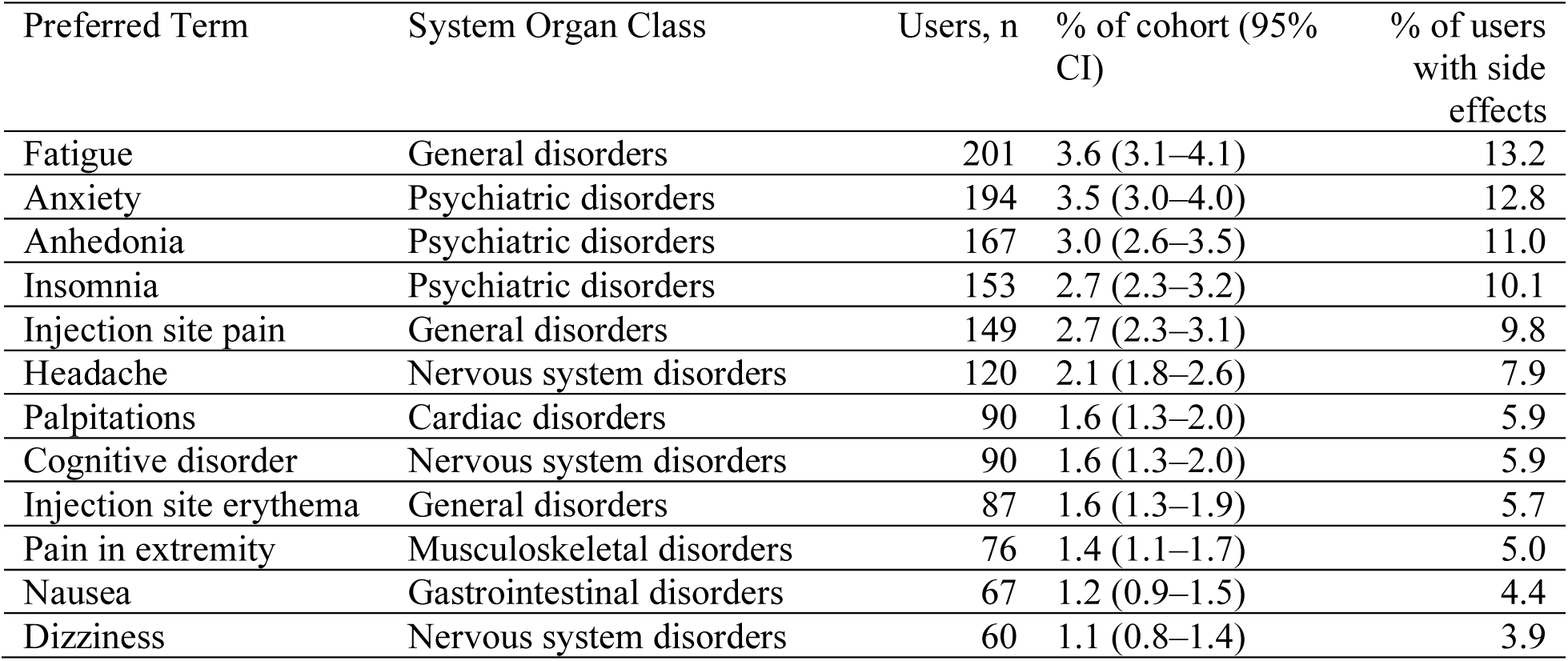
Most frequent self-reported side effects (MedDRA Preferred Terms) among 1,519 users reporting any side effect (5,588 users in the cohort).

| Preferred Term | System Organ Class | Users, n | % of cohort (95% CI) | % of users with side effects |
|---|---|---|---|---|
| Fatigue | General disorders | 201 | 3.6 (3.1–4.1) | 13.2 |
| Anxiety | Psychiatric disorders | 194 | 3.5 (3.0–4.0) | 12.8 |
| Anhedonia | Psychiatric disorders | 167 | 3.0 (2.6–3.5) | 11.0 |
| Insomnia | Psychiatric disorders | 153 | 2.7 (2.3–3.2) | 10.1 |
| Injection site pain | General disorders | 149 | 2.7 (2.3–3.1) | 9.8 |
| Headache | Nervous system disorders | 120 | 2.1 (1.8–2.6) | 7.9 |
| Palpitations | Cardiac disorders | 90 | 1.6 (1.3–2.0) | 5.9 |
| Cognitive disorder | Nervous system disorders | 90 | 1.6 (1.3–2.0) | 5.9 |
| Injection site erythema | General disorders | 87 | 1.6 (1.3–1.9) | 5.7 |
| Pain in extremity | Musculoskeletal disorders | 76 | 1.4 (1.1–1.7) | 5.0 |
| Nausea | Gastrointestinal disorders | 67 | 1.2 (0.9–1.5) | 4.4 |
| Dizziness | Nervous system disorders | 60 | 1.1 (0.8–1.4) | 3.9 |

The profile was similar after excluding the 21.6% of users reporting co-use of the drugs listed in the Methods (11.6% with a psychiatric or nervous system effect versus 13.4% overall), and fatigue, anxiety and anhedonia remained among the four most common PTs in every subgroup, including users of BPC-157 without TB-500 (Supplementary Table S7). Including 693 users who described only an unspecified injury changed no main result by more than 1.3 percentage points (Supplementary Table S11).

In the negative control, users of BPC-157 and/or TB-500 alone (n=9,065) reported side effects about as often as users of KPV, MOTS-c, Epitalon or Semax alone (26.1% versus 20.2% to 27.6%), but anhedonia was described by 10.2% of BPC-157/TB-500 users reporting side effects versus 1.3% of other peptide users (risk ratio 7.7, 95% CI 3.4–17.1; odds ratio adjusted for posting volume 6.7, 2.9–15.1) and 2.9% of users of the nootropic Semax. The excess was similar among BPC-157/TB-500 users with no musculoskeletal use (10.2%) or using the peptides for gut problems (12.9%), and users of BPC-157/TB-500 plus another peptide attributed anhedonia to BPC-157/TB-500 (76 users) far more often than to the other peptide (11). Combining anhedonia with depression, depressed mood and apathy reduced the excess (12.6% versus 3.8%; risk ratio 3.3, 2.1–5.4) to a level similar to Semax (9.6%). Fatigue (11.7% versus 15.8%) and insomnia (8.0% versus 8.0%) were not more common with BPC-157/TB-500, and anxiety was modestly more common (13.0% versus 8.0%) (Supplementary Table S13).

Physician-involved users reported side effects more often than other users (36.3% versus 26.8%; odds ratio 1.55, 95% CI 1.17–2.06), but they also posted more, and the difference was not significant after adjustment for posting volume (adjusted odds ratio 1.14, 0.83–1.55). Physician involvement did not change significantly over time (2.8% to 4.7% by year; P=0.09). The median BPC-157 dose among new users was 250 to 300 µg from 2021 to 2024 and 500 µg in 2025, when 50.9% of new users stating a dose took 500 µg or more, compared with 28.1% in 2021 (Supplementary Table S8). Users taking 500 µg or more reported side effects less often than those taking less (29.1% versus 38.6% below 300 µg and 40.9% at 300 to 499 µg; odds ratio adjusted for posting volume and year 0.64, 0.53–0.78). New users increased from 471 in 2021 to 1,883 in 2025 (Figure 4), in step with the number of people posting about any study peptide (2,258 to 10,606), so the share of active posters describing musculoskeletal use was stable at 17.8 to 20.9 per 100; the 2021 count also includes users active before the archive began. Among users reporting side effects, the proportion describing anhedonia varied by year (8.7% to 15.6%; P=0.017) without a consistent upward trend.

**Figure 4.**
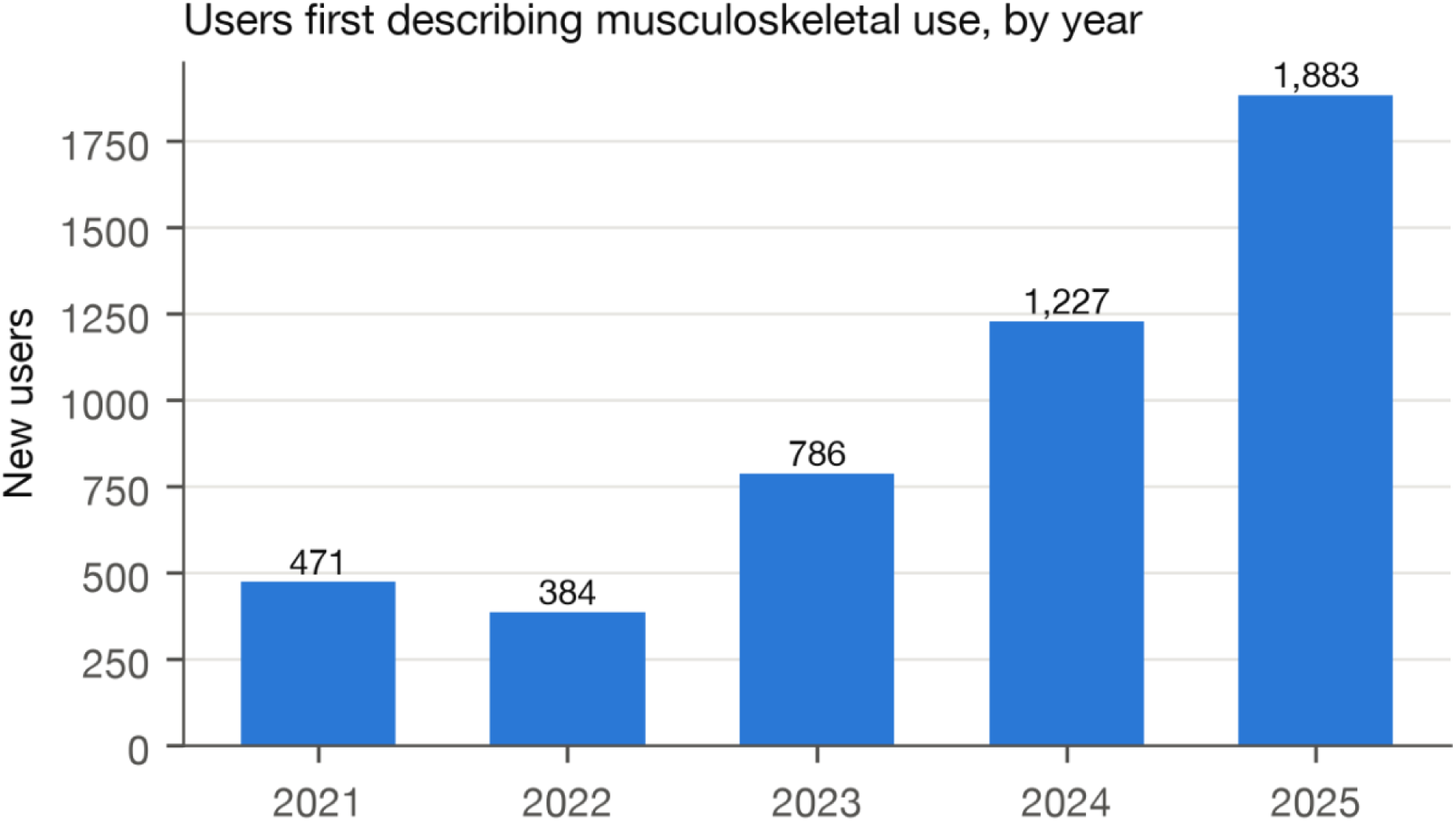
Users first describing musculoskeletal use of BPC-157 or TB-500, by year, 2021 to 2025. Detailed methods, Supplementary Tables S1 to S13, and the STROBE^30^ checklist are in the Supplementary Appendix.

## Discussion

In this analysis of 92,254 Reddit posts and comments, more than 5,500 users described using BPC-157 and/or TB-500 for a musculoskeletal injury, most often chronic tendinopathy of the shoulder, knee or elbow, often alongside physical therapy or surgery and rarely under physician supervision. Most bought their peptides from research-chemical vendors, doses rose over time, and 27% reported side effects, among which anhedonia was specific to these peptides. To our knowledge, this is the first systematic description of how these peptides are used for musculoskeletal injury outside research settings.

The injuries users reported treating differ in important ways from those modeled in preclinical studies, which created acute injuries surgically in healthy rats and began treatment at or shortly after injury.^7–10^ Chronic tendinopathy is a degenerative condition with different biology from an acute surgical wound, and whether a peptide that speeds repair of such a wound in rats affects it in people has never been tested. Because chronic tendon pain fluctuates and often improves with time, rehabilitation and the concurrent treatments many users received, the 79% who reported improvement cannot be taken as evidence that the peptides worked.

Users’ doses have no empirical basis in humans. Because drug exposure scales with body-surface area rather than weight, the 10 µg/kg rat regimen corresponds to about 1.6 µg/kg in humans, or roughly 140 µg for an 85 kg adult.^24^ This conversion guides starting doses in first-in-human trials and does not account for route, frequency or pharmacokinetics, none of which have been characterized in humans, so it does not identify a safe or effective dose. Doses are also rising: the median among new users rose from 300 µg in 2024 to 500 µg in 2025. Higher doses were not associated with more reported side effects, but this cross-sectional comparison cannot exclude harm: users who tolerated the peptides may have increased their dose while those with side effects reduced or stopped it. Most users also injected subcutaneously away from the injury, assuming that the peptide reaches injured tissue through the circulation, which has not been tested in people.

Nine in ten of those naming a source bought from research-chemical vendors, whose products are not approved by the FDA, are labeled not for human use and are not made to pharmaceutical standards,^1,4^ so their identity, purity, sterility and content are unknown. Only one in 25 users described a physician prescribing or administering the peptide. Users may simply not have mentioned such involvement. The FDA advisory committee’s recommendation to permit pharmacy compounding^19^ could improve product quality but would not provide evidence of efficacy or an evidence-based dose, and in our data physician involvement was not associated with fewer side effects. In addition, 5.7% of users were competitive athletes, for whom any use is an anti-doping violation.^18^

We also identified a side-effect profile not described in the limited human studies of these peptides, which were small, uncontrolled and not designed to detect uncommon effects.^12,13^ Fewer users reported side effects than Reddit users of semaglutide and tirzepatide in a similar analysis (43.5%),^21^ although this may reflect posting behavior as much as tolerability, and the profile differed: in place of the gastrointestinal symptoms that dominate reports for glucagon-like peptide-1 receptor agonists, reports were dominated by fatigue, anxiety, anhedonia and insomnia, although BPC-157 is derived from a gastric protein.^25^ This pattern is consistent with a non-peer-reviewed analysis of a single community.^26^

Several findings suggest that the type of person using these peptides does not fully explain the anhedonia signal. It persisted after excluding users of co-administered agents most likely to cause neuropsychiatric symptoms, was about eight times as common as with four other peptides discussed in the same communities, and did not depend on whether the peptides were used for an injury. Anhedonia was also reported by only 0.7% of Reddit users describing side effects of semaglutide or tirzepatide.^21^ BPC-157 interacts with central dopaminergic and serotonergic systems in rodents,^25,27^ which could plausibly affect reward, but its effects in rat models of depression and anxiety were antidepressant-like and anxiolytic-like,^25^ so no mechanism for anhedonia has been established. Reporting increases after a suspected effect is publicized,^28^ and a narrative attached to one peptide could prompt users to attribute their symptoms to it, which a negative-control comparison cannot exclude; the smaller excess for low-mood terms combined suggests that part of the signal reflects the words users choose. The proportion describing anhedonia did not rise consistently over time, which argues against, but cannot exclude, earlier posts prompting later reports. Nocebo effects are also possible, and misattribution of the low mood that accompanies chronic pain is less likely given that anhedonia was as common among users without an injury.

Serious events were rare (0.5% of users), and their reporting cannot establish causality, but reports of new or progressing cancers merit attention. BPC-157 promotes angiogenesis through vascular endothelial growth factor receptor 2,^29^ which may contribute to healing but could in principle support tumor growth. Most cancers described were basal cell carcinomas, which are common in middle-aged adults, but with no long-term human safety data, this theoretical concern argues for caution in people with a history of cancer and for long-term surveillance.

The main strengths of this study are its scale and its subject: no trials, registries or surveillance systems capture how these peptides are used, and pseudonymous posting may lead users to describe more candidly than in a clinic what they do. Other strengths are physician validation of each extraction step and a negative-control comparison.

### Limitations

This study has limitations. Users of peptide communities may differ from all users of these peptides, and fewer than one in ten disclosed demographic characteristics. Because side effects and outcomes were volunteered rather than elicited, we cannot estimate their true frequency or establish causality; posting may favor more severe experiences, beliefs about attribution may shape reports, and product identity, dose and diagnoses could not be verified. Reported outcomes are especially vulnerable to selection, expectation, regression to the mean and concurrent treatment. Finally, each validation post was rated by one physician, agreement on specific side-effect terms was modest, and the cohort is more likely undercounted than inflated. Our findings should be treated as hypothesis-generating signals that require confirmation by surveillance systems and prospective studies.^20^

Randomized trials in chronic tendinopathy, with pharmacokinetic sampling and prespecified neuropsychiatric outcomes, are needed before any benefit or dose can be claimed, and analytical testing of products sold by research-chemical vendors would establish what users are injecting. If compounding is permitted, a registry of patients receiving compounded peptides would allow prospective follow-up, and clinicians should report suspected adverse effects to the FDA MedWatch program.

### Conclusions

This study provides the largest description to date of how BPC-157 and TB-500 are used for musculoskeletal injury. Sports medicine clinicians should ask patients with chronic tendinopathy about peptide use and counsel them on the absence of human efficacy data, unregulated supply, anti-doping status and reported side effects, including anhedonia.

## Declarations

### Ethics

Data were collected from publicly accessible Reddit posts and comments. No attempts were made to access private or password-protected content. As data were public and non-identifiable, the Colorado Multiple Institutional Review Board determined that this study was exempt from review.

### Consent

Not applicable; no participants were recruited, no identifiers are reported, and quotations are not reproduced.

### Use of artificial intelligence

Large language models were the extraction method. The authors also used large language models to edit the manuscript. The authors reviewed all output and take full responsibility for the content.

### Author contributions

JC conceived and designed the study, built the data pipeline, performed the analysis, annotated the self-use, side-effect, physician-involvement and outcome validation samples, adjudicated serious events, and wrote the manuscript. CC and CL independently reviewed the musculoskeletal validation samples. CC, CL and VW critically reviewed the manuscript. JC is the guarantor.

### Data availability statement

All data used in this study are publicly available from Reddit.com or are accessible via Arctic Shift. Due to Reddit’s Terms of Use, we are unable to share the raw data. All Reddit data in this study were used in accordance with Reddit’s Terms of Use. Code, prompts, schemas and annotation guides to reproduce the analyses are available from the corresponding author on reasonable request.

