## Supplementary Appendix for "Real-World Use, Dosing, and Reported Side Effects of BPC-157 and TB-500 for Musculoskeletal Injury: An Analysis of Online Communities"

**Contents**


- S1. Data source and sampling frame
- S2. Variable definitions and user-level aggregation
- S3. Side-effect coding
- S4. Statistical analysis
- Supplementary tables S1 to S13
- STROBE checklist


**S1. Data source and sampling frame**

**Source.** Posts and comments were retrieved from the Arctic Shift archive of Reddit (https://arctic-shift.photon-reddit.com) through its public API between 16 and 24 September 2026. We retrieved every post and comment, without keyword filtering on the server, from the six Reddit communities dedicated to peptide use: r/Peptides, r/bpc_157, r/peptidesource, r/PeptideGuide, r/TB500, and r/Semax. The study window was 1 January 2021 to 31 August 2026. r/bpc_157 was created in mid-2021, and earlier years contribute few texts.

**Communities not analyzed.** Three large general communities (r/Biohackers, r/Nootropics, r/moreplatesmoredates) were retrieved with a keyword filter. They were excluded from the analytic sample because their coverage at the time of analysis spanned only two months, and including them would have made the sampling frame inconsistent over time. Sixteen sport and injury communities were also retrieved with a keyword filter and were not analyzed.

**Quality filters.** Following Sehgal et al., we excluded:

- deleted or removed texts
- texts by deleted accounts, AutoModerator, or accounts whose names contain "bot"
- texts of fewer than 10 words
- texts with no space in the first 2,048 characters
- texts in which fewer than 50% of characters were letters

Duplicates by Reddit ID were removed. A post's title and body were analyzed together as one text.

**Peptide mention filter.** We retained texts containing at least one alias of a study peptide as a whole word or phrase, case-insensitive (Table S1). The pipeline was built for six peptides that an FDA advisory committee considered in July 2026, and the self-use step classified use of all six. This paper analyzes BPC-157 and TB-500 only.

**Unit of analysis.** The unit of analysis was the Reddit user (pseudonymous account). A user entered the cohort if any of their texts was classified as describing personal use of BPC-157 and/or TB-500 for a musculoskeletal problem and at least one such text named a specific body region or tissue (S2).

**Table S1. Search terms (whole word or phrase, case-insensitive).**

| Peptide | Aliases |
|---|---|
| BPC-157 | bpc-157, bpc 157, bpc157, bpc, body protection compound, body protective compound, pentadecapeptide, bcp-157, bpc-175 |
| TB-500 | tb-500, tb 500, tb500, thymosin beta-4, thymosin beta 4, thymosin b4, tb-4, tb4, tβ4 |
| Combination slang (implies BPC-157 and TB-500) | wolverine stack, wolverine protocol, glow stack, glow protocol |

Aliases for the other study peptides (KPV, MOTS-c, Epitalon, Semax) are in config/peptides.yaml; texts mentioning only those peptides were classified in the self-use step but do not contribute to this analysis.

### S2. Variable definitions and user-level aggregation

Users could contribute several texts. User-level variables were derived from all of a user's musculoskeletal-use texts as follows.

- **Peptides, injury region, tissue, chronicity, diagnoses, route, injection site, source, and concurrent treatments.** The union across the user's texts. Categories are therefore not mutually exclusive, and percentages can sum to more than 100.
- **Injury location and tissue (re-extraction).** When the original extraction was checked against the text, its body-region labels proved unreliable for some regions, although they were reliable for others. We counted how often each label's text or diagnosis named that body part. This held for 84–96% of labels for the knee, shoulder, ankle and foot, wrist and hand, lumbar spine, hip, and elbow, but for only 19–38% of the cervical spine, chest, thigh, and calf and Achilles labels. Posts labeled "cervical spine" mostly described the low back, posts labeled "chest" mostly the neck, and "calf and Achilles" served as a catch-all.

  We therefore re-extracted the injuries from all musculoskeletal-use texts with a separate prompt (prompts/msk_injury.md). It used plain-language location labels and required the model to quote the words naming each location and tissue. A location or tissue without a supporting quote was set to unspecified. The re-extraction covered 12,156 texts; 1 texts that it did not return kept their original injuries. The fix was designed from the cohort-wide pattern, before it was compared with the reviewers' labels.
- **Musculoskeletal definition.** The extraction prompt and the reviewer guide counted use for an injury or musculoskeletal pain even when the author did not name the body part. Physician review found that unspecified mentions of an "injury" were often not judged musculoskeletal. Against the reviewers' labels, the model's precision for musculoskeletal use was 78.1% when these texts were counted and 96.1% when a named region or tissue was required. The analytic cohort therefore requires a specific body region or tissue (not

"unspecified" or "multiple") in at least one musculoskeletal-use text. This definition was adopted after the physician review, and the broader definition is reported as a sensitivity analysis (Table S11).

- **Combined use.** The user used both BPC-157 and TB-500 in any text.
- **Named diagnoses.** Free-text diagnoses were grouped into clinical categories with keyword rules, listed in analysis/msk_paper.py.
- **Source among users stating a source.** The denominator is users with at least one known source. "Compounding pharmacy or clinic" combines compounding pharmacy and clinic-prescribed sources.
- **Injection site among users stating a site.** The denominator is users with at least one known site, excluding oral or nasal use.
- **Physician involvement.** Any text describing a clinician prescribing, supervising, or administering the peptide, including clinic injection. The model also labeled explicit self-direction, but validation work showed that it applied this label where a reader would judge the matter unstated. We therefore report only the presence or absence of described physician involvement.
- **Reported outcome.** The verdict in the user's most recent text that stated one. The denominator excludes users who never stated an outcome.
- **Time to perceived effect.** Parsed from free text to days using the first stated quantity and unit. Weeks were converted at 7 days and months at 30 days. Number words were converted to numbers, with "a few" taken as 3 and "several" as 4. "Immediately" and "overnight" were taken as 1 day. Of 2,790 users who stated a time, 2,243 statements could be parsed.

- **Dose.** The median of the user's unambiguous per-administration doses, in micrograms, for each peptide. Values below 10 μg or above 20,000 μg were treated as unit or parsing errors and excluded: 9 users for BPC-157 and 12 for TB-500. Doses per kilogram assumed a body weight of 85 kg. Among the 180 BPC-157 users who stated their weight, the median dose per kilogram of stated weight was 4.2 μg/kg (IQR 2.8 to 5.5), similar to the 4.3 μg/kg estimate under the assumption.
- **Athlete status.** Competitive only if the author mentioned competing, a team, league, meet, race, fight, or season and named or clearly implied the sport. Sports were grouped into families.
- **Demographics.** Sex and age as the author stated them about themselves in any self-use text, extracted in the self-use step. Sex was accepted only as an unambiguous male or female statement. Age was accepted as a number from 16 to 89 or a decade ("mid 40s" as 45, "early 40s" as 42, "late 40s" as 48). The user's modal sex and median age were used.
- **Co-use of potentially confounding substances.** Detected by regular expression in the concomitant substances extracted from all of the user's self-use texts. There were four classes:
    - incretin-based drugs (semaglutide, tirzepatide, retatrutide, cagrilintide, GLP-1)
    - androgens, anabolic steroids, selective androgen receptor modulators, and related agents (testosterone, TRT, oxandrolone, nandrolone, trenbolone, SARMs, enclomiphene, clomiphene, hCG)
    - growth hormone and its secretagogues (HGH, ipamorelin, CJC-1295, tesamorelin, sermorelin, MK-677, GHRPs)
    - stimulants (amphetamines, modafinil, methylphenidate, nicotine, ephedrine, clenbuterol)
- **Year of entry.** The year of the user's first musculoskeletal-use text.

### S3. Side-effect coding

**Attribution.** Effects from any of a cohort member's self-use texts were included when the author attributed them to BPC-157, TB-500, the combination, or an unspecified study peptide. Effects attributed to other study peptides (KPV, MOTS-c, Epitalon, Semax) were excluded.

**MedDRA normalization.** We did not have a MedDRA licence. Model-assigned Preferred Term strings were normalized to canonical spellings, and each was assigned its primary System Organ Class from a curated lookup of 156 common Preferred Terms (resources/pt_soc_fallback.csv). The lookup covered 71.9% of side-effect mentions in the cohort. The remainder kept the model-assigned SOC, which was constrained to the official list. Terms were not verified against the MedDRA dictionary, and synonyms may be split across terms.

**Neuropsychiatric effects.** Any Preferred Term in the Psychiatric disorders or Nervous system disorders SOCs.

**Severity, seriousness, dechallenge, and rechallenge.**

- Severity follows the author's description:
  - mild: tolerable or brief
  - moderate: interfered with daily life or prompted a dose change
  - severe: led to stopping or medical care, or was described as severe
- Serious follows the regulatory definition. It requires death, a life-threatening event, hospitalization or emergency department visit, persistent or significant disability, a new or growing cancer, or a congenital anomaly.
- A positive rechallenge requires an effect that resolved on stopping and recurred on restarting.

Model-flagged seriousness and rechallenge were treated as screening flags pending physician review.

### S4. Statistical analysis

- **Descriptive statistics.** Proportions are reported with 95% Wilson score intervals, and continuous variables as medians with interquartile ranges.
- **Side-effect frequencies.** Reported per user: once per user per Preferred Term, and once per user per SOC. The denominator is either the cohort or users reporting any effect, as labeled.

- **Physician involvement and side effects.** Compared by logistic regression, crude and adjusted for the natural logarithm of the user's number of self-use texts. The number of texts is a measure of reporting intensity.
- **Reported outcomes by peptide.** Among users stating an outcome, improvement was reported by 77.5% of those using BPC-157 alone, 80.6% of those using both peptides, and 71.7% of those using TB-500 alone (n=60).
- **Validation estimates.** Estimates from the stratified validation sample were weighted by stratum size.
- **Trends.** New users by year of entry were described for complete years (2021 to 2025). Physician involvement across years was tested with a chi-square test.
- **Sensitivity analyses of the side-effect profile.** The analysis was repeated:
    - excluding injection-site terms
    - excluding users who reported co-use of any confounding class
    - restricted to BPC-157 without TB-500
    - restricted to BPC-157 with TB-500
    - excluding users who reported any other study peptide
    - under the broader musculoskeletal definition, which was also applied to the main characteristics (Table S11)
- **Inference.** All tests were two-sided and exploratory, and no adjustment was made for multiple comparisons.
- **Software.** Python 3.13 with pandas, statsmodels, SciPy, and matplotlib.

The single analysis script (analysis/msk_paper.py) regenerates every number, table, and figure.

### Supplementary tables

**Table S2. Fields extracted by each language-model stage.** Enumerated fields were constrained to the listed values by the JSON schema.

| Stage | Field | Values |
|---|---|---|
| 1. Self-use | Author's use | current, past, planning, none, unclear |
| | Peptides used, each with dose, route, frequency, duration, source, vendor | route: subcutaneous, intramuscular, oral, nasal, topical, intravenous, unknown; source: research vendor, compounding pharmacy, clinic prescribed, unknown |
| | Concomitant substances; indication; age; sex | free text as written |
| | Mentions own unwanted effect | true/false |
| Musculoskeletal | Musculoskeletal use | true/false |
| | Injuries: region; tissue; diagnosis; chronicity; duration before starting | 13 regions; 10 tissues; free text; acute, chronic, post-surgical, unknown |
| | Regimens: peptide; dose as written; dose (µg per administration); frequency; route; | site: near the injury, intra-articular, away from the injury, not applicable, unknown |

| Stage | Field | Values |
|---|---|---|
| | injection site; cycle length; source | |
| | Combined BPC-157 and TB-500 | true/false |
| | Concurrent treatments | physical therapy, surgery, PRP, corticosteroid injection, NSAID, rest or offloading, other peptide, testosterone or anabolic steroid, collagen or supplement, other |
| | Physician involvement | physician prescribed, clinic administered, self-directed, unknown |
| | Reported outcome; outcome quote | improved, no change, worse, mixed, too early, not reported |
| | Time to perceived effect; return to sport; athlete status; sport; body weight | return to sport: yes, partial, no, not reported; athlete: competitive, recreational, not stated |
| Injury re-extraction (replaces the musculoskeletal stage's injury fields) | Per injury: location quote; location; tissue quote; tissue; diagnosis; chronicity | location: neck, mid or lower back, shoulder, elbow or forearm, wrist or hand, chest or ribs, hip, groin or pelvis, thigh, knee, calf, Achilles or shin, ankle or foot, multiple, other, not stated; a location or tissue without a supporting quote is set to not stated |
| Side effects | Per effect: verbatim quote; peptide blamed; MedDRA PT; SOC; severity; serious; onset; dechallenge; rechallenge | SOC: 27 official names; severity: mild, moderate, severe, unknown; dechallenge: resolved, persisted, not stopped, unknown; rechallenge: recurred, did not recur, not restarted, unknown |

**Table S3. Extended characteristics of 5,588 users describing musculoskeletal use of BPC-157 and/or TB-500.**

| Characteristic | Users, n | % (95% CI) |
|---|---:|---:|
| **Peptides** | | |
| BPC-157 | 5,507 | 98.6 (98.2–98.8) |
| TB-500 | 3,362 | 60.2 (58.9–61.4) |
| Both | 3,281 | 58.7 (57.4–60.0) |
| **Demographics, where stated** | | |
| Male (of 373 stating sex) | 260 | 69.7 (64.9–74.1) |
| Age, median (IQR), years (n=455) | 42 (33–50) | |
| **Injury chronicity** | | |
| Chronic (≥6 weeks) | 2,666 | 47.7 (46.4–49.0) |
| Post-surgical | 776 | 13.9 (13.0–14.8) |
| Acute (<6 weeks) | 703 | 12.6 (11.7–13.5) |

| Characteristic | Users, n | % (95% CI) |
|---|---|---|
| **Body region** | | |
| Shoulder | 2,125 | 38.0 (36.8–39.3) |
| Knee | 1,354 | 24.2 (23.1–25.4) |
| Elbow | 1,005 | 18.0 (17.0–19.0) |
| Unspecified | 984 | 17.6 (16.6–18.6) |
| Thoracolumbar spine | 959 | 17.2 (16.2–18.2) |
| Multiple | 528 | 9.4 (8.7–10.2) |
| Ankle and foot | 483 | 8.6 (7.9–9.4) |
| Hip and groin | 423 | 7.6 (6.9–8.3) |
| Wrist and hand | 414 | 7.4 (6.8–8.1) |
| Calf and Achilles | 317 | 5.7 (5.1–6.3) |
| Cervical spine | 260 | 4.7 (4.1–5.2) |
| Other | 227 | 4.1 (3.6–4.6) |
| Thigh | 216 | 3.9 (3.4–4.4) |
| Chest and pectoral | 165 | 3.0 (2.5–3.4) |
| **Tissue** | | |
| Tendon | 2,357 | 42.2 (40.9–43.5) |
| Unspecified | 2,321 | 41.5 (40.2–42.8) |
| Cartilage or joint | 1,232 | 22.0 (21.0–23.2) |
| Multiple | 828 | 14.8 (13.9–15.8) |
| Muscle | 588 | 10.5 (9.7–11.4) |
| Ligament | 481 | 8.6 (7.9–9.4) |
| Nerve | 433 | 7.7 (7.1–8.5) |
| Intervertebral disc | 401 | 7.2 (6.5–7.9) |
| Bone | 281 | 5.0 (4.5–5.6) |
| Fascia | 208 | 3.7 (3.3–4.3) |
| **Named diagnosis (grouped)** | | |
| Tendinopathy, site unspecified | 859 | 15.4 (14.5–16.3) |
| Rotator cuff tendinopathy or tear | 370 | 6.6 (6.0–7.3) |
| Disc herniation or bulge | 322 | 5.8 (5.2–6.4) |
| Lateral epicondylopathy (tennis elbow) | 285 | 5.1 (4.6–5.7) |
| Osteoarthritis or arthritis | 192 | 3.4 (3.0–3.9) |
| Labral tear | 190 | 3.4 (3.0–3.9) |
| Meniscal tear | 185 | 3.3 (2.9–3.8) |
| ACL injury | 152 | 2.7 (2.3–3.2) |
| Medial epicondylopathy (golfer's elbow) | 127 | 2.3 (1.9–2.7) |
| Achilles tendinopathy or rupture | 91 | 1.6 (1.3–2.0) |
| Sciatica or radiculopathy | 60 | 1.1 (0.8–1.4) |
| MCL injury | 58 | 1.0 (0.8–1.3) |
| **Concurrent treatment** | | |
| Physical therapy | 2,048 | 36.6 (35.4–37.9) |
| Rest or offloading | 1,076 | 19.3 (18.2–20.3) |
| Surgery | 974 | 17.4 (16.5–18.4) |
| Other | 882 | 15.8 (14.9–16.8) |
| Other peptide | 824 | 14.7 (13.8–15.7) |
| Corticosteroid injection | 410 | 7.3 (6.7–8.1) |
| NSAID | 367 | 6.6 (5.9–7.2) |
| Platelet-rich plasma | 332 | 5.9 (5.4–6.6) |

| Characteristic | Users, n | % (95% CI) |
|---|---|---|
| Testosterone or anabolic steroid | 202 | 3.6 (3.2–4.1) |
| Collagen or supplement | 115 | 2.1 (1.7–2.5) |
| **Route** | | |
| Subcutaneous | 4,263 | 76.3 (75.2–77.4) |
| Oral | 645 | 11.5 (10.7–12.4) |
| Intramuscular | 268 | 4.8 (4.3–5.4) |
| Nasal | 48 | 0.9 (0.6–1.1) |
| Intravenous | 33 | 0.6 (0.4–0.8) |
| Topical | 18 | 0.3 (0.2–0.5) |
| **Injection site (of 4,450 stating a site)** | | |
| Away from the injury | 3,844 | 86.4 (85.3–87.4) |
| Near the injury | 1,287 | 28.9 (27.6–30.3) |
| **Source (of 2,132 stating a source)** | | |
| Research-chemical vendor | 1,939 | 90.9 (89.7–92.1) |
| Compounding pharmacy or clinic | 338 | 15.9 (14.4–17.5) |
| **Oversight** | | |
| Any physician involvement described | 215 | 3.8 (3.4–4.4) |
| No physician involvement described | 5,373 | 96.2 (95.6–96.6) |
| **Co-use of potentially confounding substances** | | |
| Incretin-based drug | 371 | 6.6 (6.0–7.3) |
| Androgen, anabolic steroid or SARM | 379 | 6.8 (6.2–7.5) |
| Growth hormone or secretagogue | 779 | 13.9 (13.1–14.9) |
| Stimulant | 97 | 1.7 (1.4–2.1) |
| Any of the above | 1,208 | 21.6 (20.6–22.7) |
| **Dose per administration, median (IQR) [5th–95th percentile], µg** | | |
| BPC-157 (n=2,158) | 366 (250–500) [200–1,000] | |
| BPC-157 per kg, assuming 85 kg | 4.3 (2.9–5.9) µg/kg | |
| BPC-157 per kg of stated weight (n=180) | 4.2 (2.8–5.5) µg/kg | |
| BPC-157 at or above 10 µg/kg (rat regimen) | | 9.0 (7.9–10.3) |
| TB-500 (n=1,238) | 500 (350–2,000) [250–5,000] | |
| **Reported outcome (of 4,832 stating one)** | | |
| Improved | 3,832 | 79.3 (78.1–80.4) |
| Mixed | 94 | 1.9 (1.6–2.4) |
| No change | 616 | 12.7 (11.8–13.7) |
| Too early to tell | 97 | 2.0 (1.6–2.4) |
| Worse | 193 | 4.0 (3.5–4.6) |
| Time to perceived effect, median (IQR), days (n=2,243) | 10.5 (7–21) | |
| **Return to sport (of 855 reporting)** | | |
| Yes | 559 | 65.4 (62.1–68.5) |
| Partial | 268 | 31.3 (28.3–34.5) |
| No | 28 | 3.3 (2.3–4.7) |
| **Sport** | | |
| Competitive athlete | 321 | 5.7 (5.2–6.4) |

| Characteristic | Users, n | % (95% CI) |
|---|---|---|
| Any sport named | 853 | 15.3 (14.3–16.2) |
| Strength sports (of sport named) | 257 | 30.1 (27.1–33.3) |
| Combat sports (of sport named) | 187 | 21.9 (19.3–24.8) |
| Endurance sports (of sport named) | 107 | 12.5 (10.5–14.9) |
| Other (of sport named) | 91 | 10.7 (8.8–12.9) |
| Team ball sports (of sport named) | 89 | 10.4 (8.6–12.7) |
| Racquet and club sports (of sport named) | 82 | 9.6 (7.8–11.8) |
| Board and ski sports (of sport named) | 21 | 2.5 (1.6–3.7) |
| Climbing (of sport named) | 19 | 2.2 (1.4–3.5) |

Categories are not mutually exclusive: a user contributes to every category mentioned in any of their texts. Outcomes are self-reported and are not evidence of efficacy.

**Table S4. Self-reported side effects among 5,588 users, by System Organ Class and Preferred Term.**

1,519 users (27.2%; 95% CI 26.0–28.4) reported 5,639 side-effect mentions covering 806 distinct Preferred Terms (mean 2.8 terms per reporting user). 746 users (13.4%; 12.5–14.3) reported at least one psychiatric or nervous system effect.

| System Organ Class | Users, n | % of users reporting any effect (95% CI) |
|---|---|---|
| General disorders and administration site conditions | 665 | 43.8 (41.3–46.3) |
| Psychiatric disorders | 504 | 33.2 (30.9–35.6) |
| Nervous system disorders | 430 | 28.3 (26.1–30.6) |
| Musculoskeletal and connective tissue disorders | 277 | 18.2 (16.4–20.3) |
| Gastrointestinal disorders | 234 | 15.4 (13.7–17.3) |
| Skin and subcutaneous tissue disorders | 158 | 10.4 (9.0–12.0) |
| Cardiac disorders | 127 | 8.4 (7.1–9.9) |
| Vascular disorders | 88 | 5.8 (4.7–7.1) |
| Metabolism and nutrition disorders | 76 | 5.0 (4.0–6.2) |
| Eye disorders | 61 | 4.0 (3.1–5.1) |

| Preferred Term | Users, n | % of cohort (95% CI) | % of users reporting any effect |
|---|---|---|---|
| Fatigue | 201 | 3.6 (3.1–4.1) | 13.2 |
| Anxiety | 194 | 3.5 (3.0–4.0) | 12.8 |
| Anhedonia | 167 | 3.0 (2.6–3.5) | 11.0 |

| Preferred Term | Users, n | % of cohort (95% CI) | % of users reporting any effect |
|---|---|---|---|
| Insomnia | 153 | 2.7 (2.3–3.2) | 10.1 |
| Injection site pain | 149 | 2.7 (2.3–3.1) | 9.8 |
| Headache | 120 | 2.1 (1.8–2.6) | 7.9 |
| Palpitations | 90 | 1.6 (1.3–2.0) | 5.9 |
| Cognitive disorder | 90 | 1.6 (1.3–2.0) | 5.9 |
| Injection site erythema | 87 | 1.6 (1.3–1.9) | 5.7 |
| Pain in extremity | 76 | 1.4 (1.1–1.7) | 5.0 |
| Nausea | 67 | 1.2 (0.9–1.5) | 4.4 |
| Dizziness | 60 | 1.1 (0.8–1.4) | 3.9 |
| Pruritus | 58 | 1.0 (0.8–1.3) | 3.8 |
| Injection site reaction | 56 | 1.0 (0.8–1.3) | 3.7 |
| Lethargy | 55 | 1.0 (0.8–1.3) | 3.6 |

| Characteristic of user–term pairs (model-extracted, before validation) | n | % |
|---|---|---|
| Severity: mild | 1,329 | 31.2 |
| Severity: moderate | 2,335 | 54.8 |
| Severity: severe | 561 | 13.2 |
| Severity: unknown | 38 | 0.9 |
| Mentions with resolution on stopping (positive dechallenge) | 1,520 | |
| Mentions with recurrence on restarting (positive rechallenge); users | 251; 106 | not validated |
| Mentions flagged serious by the model; texts; users | 173; 119; 59 | see Table S12 |

Most frequent Preferred Terms among model-flagged serious mentions, before physician review:

Neoplasm malignant 32; Anaphylactic reaction 9; Anhedonia 6; Autoimmune disorder 6;

Anxiety 6; Panic attack 4; Abdominal pain 4; Appendicitis 4; Depression 3; Tachycardia 3.

Physician-confirmed serious events are in Table S12.

**Table S5. Users reporting each combination of injury region and tissue (data for Figure 2).**

A user may contribute to more than one cell.

| Region Tissue | Bone | Cartilage or joint | Intervertebral disc | Fascia | Ligament | Multiple | Muscle | Nerve | Tendon | Unspecified |
|---|---|---|---|---|---|---|---|---|---|---|

| Region Tissue | Bone | Cartilage or joint | Intervertebral disc | Fascia | Ligament | Multiple | Muscle | Nerve | Tendon | Unspecified |
|---|---|---|---|---|---|---|---|---|---|---|
| Ankle and foot | 70 | 51 | 0 | 56 | 87 | 60 | 2 | 19 | 62 | 148 |
| Chest and pectoral | 17 | 10 | 0 | 16 | 2 | 6 | 83 | 3 | 15 | 24 |
| Elbow | 7 | 30 | 0 | 3 | 17 | 29 | 42 | 41 | 774 | 200 |
| Hip and groin | 17 | 136 | 3 | 15 | 11 | 63 | 74 | 19 | 57 | 142 |
| Knee | 25 | 478 | 1 | 14 | 222 | 152 | 7 | 7 | 288 | 493 |
| Calf and Achilles | 39 | 5 | 0 | 25 | 4 | 12 | 41 | 8 | 180 | 35 |
| Multiple | 3 | 209 | 1 | 1 | 17 | 173 | 34 | 19 | 88 | 73 |
| Other | 9 | 13 | 0 | 57 | 5 | 50 | 10 | 25 | 18 | 68 |
| Shoulder | 36 | 364 | 0 | 5 | 78 | 224 | 99 | 24 | 989 | 872 |
| Cervical spine | 11 | 24 | 67 | 0 | 4 | 25 | 22 | 65 | 3 | 91 |
| Thoracolumbar spine | 33 | 70 | 351 | 2 | 12 | 87 | 83 | 183 | 19 | 395 |
| Thigh | 6 | 0 | 0 | 8 | 1 | 10 | 122 | 6 | 58 | 31 |
| Unspecified | 15 | 18 | 0 | 13 | 27 | 55 | 59 | 54 | 260 | 662 |
| Wrist and hand | 37 | 76 | 0 | 8 | 35 | 43 | 8 | 42 | 117 | 136 |

**Table S6. Side effects and reported outcomes by described physician involvement.**

| | Physician involvement described | No physician involvement described |
|---|---:|---:|
| Users, n | 215 | 5,373 |
| Any side effect, % (95% CI) | 36.3 (30.1–42.9) | 26.8 (25.7–28.0) |
| Improved, % of those stating an outcome (95% CI) | 76.9 (70.1–82.5) | 79.4 (78.2–80.5) |
| Median BPC-157 dose, µg | 375 | 357 |
| Median musculoskeletal texts per user | 2 | 1 |

Odds ratio for any side effect, physician involvement versus none described: crude 1.55 (95% CI 1.17–2.06); adjusted for log number of self-use texts 1.14 (0.83–1.55; P=0.42). The comparison group is every other user in the cohort; an explicit self-directed label was not used because it agreed with physician review no better than chance (S2). This comparison is exploratory.

**Table S7. Sensitivity analyses of the side-effect profile.** Top terms are percentages of users reporting any side effect in that group.

| Analysis | Users | Any side effect, % (95% CI) | Any psychiatric or nervous system effect, % (95% CI) | Top terms (% of reporters) |
|---|---|---|---|---|
| All users (primary analysis) | 5,588 | 27.2 (26.0–28.4) | 13.4 (12.5–14.3) | Fatigue 13.2; Anxiety 12.8; Anhedonia 11.0; Insomnia 10.1; Injection site pain 9.8 |
| Excluding injection-site terms | 5,588 | 24.9 (23.8–26.0) | 13.4 (12.5–14.3) | Fatigue 14.5; Anxiety 14.0; Anhedonia 12.0; Insomnia 11.0; Headache 8.6 |
| No reported incretin, androgen/SARM, GH/secretagogue or stimulant co-use | 4,380 | 24.0 (22.8–25.3) | 11.6 (10.6–12.5) | Fatigue 12.6; Anxiety 12.2; Anhedonia 9.9; Insomnia 9.5; Injection site pain 8.6 |
| BPC-157 without TB-500 | 2,226 | 26.0 (24.2–27.8) | 13.3 (12.0–14.8) | Anxiety 13.3; Fatigue 12.8; Anhedonia 10.7; Insomnia 10.0; Headache 9.0 |
| BPC-157 with TB-500 | 3,281 | 27.8 (26.3–29.4) | 13.3 (12.1–14.5) | Fatigue 13.0; Injection site pain 12.4; Anxiety 12.3; Anhedonia 11.1; Insomnia 10.3 |
| No other study peptide reported | 5,019 | 26.2 (25.0–27.4) | 12.7 (11.8–13.7) | Anxiety 12.9; Fatigue 12.9; Anhedonia 10.6; Insomnia 9.5; Injection site pain 9.4 |

| Analysis | Users | Any side effect, % (95% CI) | Any psychiatric or nervous system effect, % (95% CI) | Top terms (% of reporters) |
|---|---|---|---|---|
| Broad musculoskeletal definition (any injury, including unspecified) | 6,281 | 26.6 (25.5–27.7) | 13.1 (12.2–13.9) | Fatigue 13.3; Anxiety 12.9; Anhedonia 11.2; Injection site pain 10.3; Insomnia 10.0 |

**Table S8. Characteristics of new users by year of first musculoskeletal text, 2021–2025.** Chi-square test for physician involvement across years, P=0.09. 2026 (January to August) is omitted as an incomplete year (837 new users).

| Year | New users | Physician involvement, % | Research vendor among source stated, % | Median BPC-157 dose, µg | Any side effect, % | Oral route, % |
|---|---|---|---|---|---|---|
| 2021 | 471 | 4.2 | 91.4 | 295 | 29.7 | 8.3 |
| 2022 | 384 | 3.6 | 90.8 | 253 | 29.9 | 10.9 |
| 2023 | 786 | 4.7 | 88.9 | 250 | 32.1 | 11.6 |
| 2024 | 1,227 | 4.3 | 91.0 | 300 | 29.8 | 14.1 |
| 2025 | 1,883 | 2.8 | 93.2 | 500 | 23.8 | 12.6 |

**Table S9. Validation of model extraction against physician labels.** Musculoskeletal-stage rows compare the model with the single reviewer who rated each text (two reviewers, 50 texts each); self-use and side-effect rows compare the model with JC's blind labels, scored for BPC-157 and TB-500 only.

| Stage and field | Texts, n | Reference positives, n | Precision, % (95% CI) | Recall, % (95% CI) | F1 | Agreement with reviewer |
|---|---|---|---|---|---|---|
| Self-use: author used BPC-157 or TB-500 | 25 | 28 | 91.3 (73–98) | 75.0 (57–87) | 82.4 | single rater (JC) |
| Self-use: author described an unwanted effect of their | 25 | 6 | 85.7 (49–97) | 100.0 (61–100) | 92.3 | single rater (JC) |

| Stage and field | Texts, n | Reference positives, n | Precision, % (95% CI) | Recall, % (95% CI) | F1 | Agreement with reviewer |
|---|---|---|---|---|---|---|
| own use | | | | | | |
| Side effects: Preferred Term, exact match | 25 | 21 terms | 35.0 (18–57) | 33.3 (17–55) | 34.1 | single rater (JC) |
| Side effects: Preferred Term, same SOC | 25 | 21 terms | 60.0 (39–78) | 57.1 (37–76) | 58.5 | single rater (JC) |
| Musculo skeletal use, cohort definitio n (specific region or tissue) | 100 | 52 | 96.1 (87–99) | 94.2 (84–98) | 95.1 | κ=0.90 |
| Musculo skeletal use, broad definitio n | 100 | 52 | 78.1 (67–86) | 96.2 (87–99) | 86.2 | κ=0.68 |
| Body region of the main injury | 46/50 | | | | | 92.0 (81–97) |
| Tissue of the main injury | 32/50 | | | | | 64.0 (50–76) |
| Physicia n involvem ent described | 42/50 | | | | | 84.0 (71–92) |
| Reported outcome | 32/50 | | | | | 64.0 (50–76) |
| BPC-157 dose within 10% | 12/13 | | | | | 92.3 (67–99) |
| TB-500 | 5/7 | | | | | 71.4 (36–92) |

| Stage and field | Texts, n | Reference positives, n | Precision, % (95% CI) | Recall, % (95% CI) | F1 | Agreement with reviewer |
|---|---|---|---|---|---|---|
| dose within 10% | | | | | | |
| Serious events (user level): model flag confirmed by physician | 59 users flagged | 29 confirmed | 49.2 | not estimable | | single rater (JC) |

Region, tissue, physician involvement, outcome, and dose are scored among texts that both the model and the reviewer classified as musculoskeletal use; the denominator is the number of those texts the reviewer labelled for that field.

**Table S10. Study flow and contribution by community.**

| Step | Texts | Users |
|---|---|---|
| Texts naming a study peptide after quality filters | 92,254 | |
| Classified in the self-use step | 91,928 | |
| Disclosing personal use of any study peptide | 34,071 | 13,456 |
| Disclosing personal use of BPC-157 or TB-500 | 28,199 | 11,458 |
| Passing the musculoskeletal keyword screen | 18,234 | |
| Extracted in the musculoskeletal stage | 18,207 | |
| Describing musculoskeletal use | 12,157 | 6,281 |
| Users naming a specific body region or tissue (analytic cohort) | 11,407 | 5,588 |
| Sent to the side-effect stage (self-use texts flagging an unwanted effect) | 9,233 | |

| Community | Musculoskeletal-use texts |
|---|---|

| Community | Musculoskeletal-use texts |
|---|---|
| r/Peptides | 5,901 |
| r/bpc_157 | 4,675 |
| r/Peptidesource | 605 |
| r/PeptideGuide | 189 |
| r/TB500 | 36 |
| r/Semax | 1 |

Self-use classifications of all texts: none 38,954; current 31,871; planning 14,681; unclear 3,607; past 2,815.

**Table S11. Main results under the cohort (strict) and broad musculoskeletal definitions.** The cohort definition requires a specific body region or tissue named in at least one text (5,588 users); the broad definition also counts users who described only an unspecified injury (6,281 users). Values are % (95% CI) unless stated.

| Characteristic | Primary (strict) definition | Broad definition |
|---|---|---|
| Users | 5,588 | 6,281 |
| Chronic, % of stating | 75.2 (73.7–76.6) | 74.1 (72.7–75.5) |
| Tendon, % of stating | 50.4 (48.9–51.8) | 49.1 (47.7–50.5) |
| Shoulder, % of stating | 38.9 (37.6–40.2) | 38.1 (36.8–39.3) |
| Knee, % of stating | 24.8 (23.7–26.0) | 24.2 (23.1–25.4) |
| Elbow, % of stating | 18.4 (17.4–19.5) | 18.0 (17.0–19.0) |
| Lumbar or thoracic spine, % of stating | 17.6 (16.6–18.6) | 17.2 (16.2–18.2) |
| Median BPC-157 dose, µg | 365.5 | 375 |
| Research vendor among source stated, % | 90.9 (89.7–92.1) | 90.9 (89.7–92.0) |
| Physician involvement described, % | 3.8 (3.4–4.4) | 3.7 (3.3–4.2) |
| Improved, % of outcome stated | 79.3 (78.1–80.4) | 79.3 (78.2–80.4) |
| Any side effect, % | 27.2 (26.0–28.4) | 26.6 (25.5–27.7) |
| Any psychiatric or nervous system effect, % | 13.4 (12.5–14.3) | 13.1 (12.2–13.9) |

**Table S12. Physician-confirmed serious events, by category.** The model flagged 173 events in 119 texts from 59 users; the physician confirmed 123 events in 29 users. Each user is counted once, in the category of their main event.

| Category | Users | Events | Examples reported by authors |
|---|---|---|---|
| New or progressing cancer | 6 | 35 | basal cell carcinoma requiring excision or Mohs surgery (4 |

| Category | Users | Events | Examples reported by authors |
|---|---|---|---|
| | | | users); appendiceal neuroendocrine tumour; metastatic gastric cancer |
| Persistent or disabling illness | 5 | 36 | multisystem neurological or autoimmune illness described as disabling; joint inflammation lasting months; lumbar vertebral collapse with two months' immobility |
| Cardiovascular emergency or event | 5 | 6 | emergency care for tachycardia (2 users, one by ambulance); chest pain with rectal bleeding; myocardial infarction; hospital assessment for suspected thrombosis |
| Gastrointestinal emergency or admission | 4 | 12 | admission for colitis; emergency endoscopy after rapid weight loss, recurring on re-exposure; admission for abdominal pain; appendicitis |
| Psychiatric emergency or admission | 4 | 12 | panic attacks requiring sedation; psychiatric admission with suicidality; emergency care for insomnia and anxiety; admission for anxiety |
| Severe allergic reaction | 3 | 13 | anaphylaxis with loss of consciousness; reactions treated with adrenaline or emergency care |
| Other emergency | 2 | 9 | emergency visits for |

| Category | Users | Events | Examples reported by authors |
|---|---|---|---|
| visit | | | unspecified or mixed reactions |

Of the 173 flagged events, 123 were confirmed serious, 28 were real but not serious, and 22 were not a serious effect of the author's own BPC-157 or TB-500 use (feared or hypothetical, another person's, or attributed to another drug).

**Table S13. Negative control: side effects among users of each study peptide alone.** Users whose self-use posts named only BPC-157 and/or TB-500, or only one other study peptide, in the same communities and processed by the same pipeline. Term percentages are among users reporting any side effect attributed to their peptide; values are % (95% CI).

| | BPC-157/TB-500 | KPV | MOTS-c | Epitalon | Semax |
|---|---|---|---|---|---|
| Users, n | 9,065 | 253 | 844 | 222 | 514 |
| Any side effect, % of users | 26.1 (25.2–27.0) | 26.1 (21.1–31.8) | 27.6 (24.7–30.7) | 20.7 (15.9–26.5) | 20.2 (17.0–23.9) |
| Users reporting side effects, n | 2,363 | 66 | 233 | 46 | 104 |
| Anhedonia | 10.2 (9.1–11.5) | 1.5 (0.3–8.1) | 0.9 (0.2–3.1) | 0.0 (0.0–7.7) | 2.9 (1.0–8.1) |
| Fatigue | 11.7 (10.5–13.1) | 10.6 (5.2–20.3) | 15.0 (11.0–20.2) | 10.9 (4.7–23.0) | 23.1 (16.0–32.0) |
| Anxiety | 13.0 (11.7–14.4) | 7.6 (3.3–16.5) | 2.6 (1.2–5.5) | 8.7 (3.4–20.3) | 20.2 (13.6–28.9) |
| Insomnia | 8.0 (7.0–9.2) | 6.1 (2.4–14.6) | 5.6 (3.3–9.3) | 23.9 (13.9–37.9) | 7.7 (3.9–14.4) |
| Headache | 8.0 (7.0–9.2) | 4.5 (1.6–12.5) | 2.1 (0.9–4.9) | 4.3 (1.2–14.5) | 11.5 (6.7–19.1) |
| Any psychiatric disorder | 32.5 (30.6–34.4) | 21.2 (13.1–32.5) | 10.7 (7.4–15.4) | 47.8 (34.1–61.9) | 49.0 (39.6–58.5) |
| Any nervous system disorder | 27.8 (26.0–29.6) | 13.6 (7.3–23.9) | 15.5 (11.4–20.6) | 39.1 (26.4–53.5) | 41.3 (32.4–51.0) |
| Any gastrointestinal disorder | 15.7 (14.3–17.3) | 18.2 (10.7–29.1) | 7.3 (4.6–11.4) | 10.9 (4.7–23.0) | 2.9 (1.0–8.1) |

Anhedonia, BPC-157/TB-500 versus the four other peptides pooled (1.3%): risk ratio 7.7 (95% CI 3.4–17.1); odds ratio among users reporting side effects adjusted for log number of self-use texts 6.7 (2.9–15.1). Anhedonia, depression, depressed mood or apathy combined: 12.6% (11.3–14.0) with BPC-157/TB-500 versus 3.8% (2.4–6.0) pooled (risk ratio 3.3, 2.1–5.4) and 9.6% (5.3–16.8) with Semax. Among BPC-157/TB-500 users reporting side effects, anhedonia was described by 10.3% (8.7–12.1) of those with musculoskeletal use, 10.2% (8.6–12.1) of those without, and 12.9% (8.4–19.5) of those using it for gut problems. Among 2,537 users of BPC-157/TB-500 and another study peptide, 76 attributed anhedonia to BPC-157 or TB-500 and 11 to another study peptide.

**STROBE checklist (cross-sectional studies)**

| Item | No. | Recommendation | Where reported |
|---|---|---|---|
| Title and abstract | 1 | Indicate design; informative, balanced summary | Title; Abstract |
| Background/rationale | 2 | Scientific background and rationale | Introduction |
| Objectives | 3 | State specific objectives | Introduction, final paragraph |
| Study design | 4 | Present key elements of design early | Methods, Data source; S1 |
| Setting | 5 | Setting, locations, periods of data collection | Methods, Data source; S1 |
| Participants | 6 | Eligibility criteria and selection | Methods, Data source; S1 (filters, unit of analysis); Figure 1 |
| Variables | 7 | Define outcomes, exposures, confounders | Methods, Language-model extraction; S2, S3; Table S2 |
| Data sources/measurement | 8 | Sources and methods of measurement; comparability | Methods, Language-model extraction and Validation; Table S9 |
| Bias | 9 | Efforts to address sources of bias | Methods, Validation and Statistical analysis; S2 (definitions); S4 (adjustment for reporting intensity, sensitivity analyses) |

| Item | No. | Recommendation | Where reported |
|---|---|---|---|
| Study size | 10 | How the study size was arrived at | All eligible texts; Figure 1 |
| Quantitative variables | 11 | Handling of quantitative variables | S2 (dose, time, age) |
| Statistical methods | 12 | Methods, subgroups, missing data, sensitivity analyses | Methods, Statistical analysis; S4; missing data handled by stating denominators (S2) |
| Participants | 13 | Numbers at each stage; reasons for non-participation; flow diagram | Results; Figure 1; Table S10 |
| Descriptive data | 14 | Characteristics; number with missing data | Table 1; Table S3 (denominators) |
| Outcome data | 15 | Numbers of outcome events | Table 2; Table S4 |
| Main results | 16 | Unadjusted and adjusted estimates with precision | Results; Tables S6 and S13 |
| Other analyses | 17 | Subgroups, interactions, sensitivity analyses | Tables S6–S8, S11 |
| Key results | 18 | Summarize key results | Discussion, first paragraph |
| Limitations | 19 | Sources of bias or imprecision | Limitations |
| Interpretation | 20 | Cautious overall interpretation | Discussion; Conclusions |
| Generalisability | 21 | External validity | Limitations; Equity, diversity and inclusion statement |
| Funding | 22 | Source of funding and role of funders | Funding statement |
